# Beyond Hypertension: Examining Variable Blood Pressure’s Role in Cognition and Brain Structure

**DOI:** 10.1101/2024.01.15.24301335

**Authors:** Cassandra Morrison, Michael D. Oliver, Farooq Kamal, Mahsa Dadar, the Alzheimer’s Disease Neuroimaging Initiative

## Abstract

**Importance:** Hypertension is a known risk factor for cognitive decline and structural brain changes in aging and dementia. In addition to high blood pressure (BP), individuals may also experience variable BP, meaning that their BP fluctuates between normal and high. It is currently unclear what the effects of variable BP are on cognition and brain structure.

**Objective:** To investigate the influence of BP on cognition and brain structure in older adults.

**Design, Setting, and Participants:** This longitudinal cohort study included data from the Rush Alzheimer’s Disease Center Research Resource Sharing Hub (RUSH) and the Alzheimer’s Disease Neuroimaging Initiative (ADNI). Participants from the two studies were included if they had BP measurements and either cognitive scores or MRI scans from at least one visit.

**Main Outcomes and Measures:** Longitudinal gray matter, white matter, white matter hyperintensity volumes, postmortem neuropathology information, as well as cognitive test scores.

**Results:** A total of 4606 participants (3429 females, mean age = 76.8) with 32776 follow-ups (mean = 7 years) from RUSH and 2114 participants (1132 females, mean age = 73.3) with 9827 follow-ups (mean = 3 years) from ADNI were included in this study. Participants were divided into one of three groups: 1) normal BP, high BP, or variable BP. Older adults with variable BP exhibited the highest rate of cognitive decline followed by high BP and then normal BP. Increased GM volume loss and WMH burden was also observed in variable BP compared to high and normal BP. With respect to post-mortem neuropathology, both variable and high BP had increased severities compared to normal BP. Importantly, results were consistent across the RUSH and ADNI participants, supporting the generalizability of the findings.

**Conclusion and Relevance:** Limited research has examined the long-term impact of variable BP on cognition and brain structure. These findings show the importance that both high and variable BP have on cognitive decline and structural brain changes. Structural damages caused by variable BP may reduce resilience to future dementia-related pathology and increased risk of dementia. Improved treatment and management of variable BP may help reduce cognitive decline in the older adult population.

## 1. Introduction

Elevated blood pressure (BP), or hypertension, is a well-established risk factor for cognitive decline and dementia.^1^ Hypertension is not only associated with cognitive decline, but also contributes to structural changes in the brain. For example, hypertension is associated with increased white matter hyperintensity (WMH)^2–4^ burden and increased neurodegeneration^5,6^ both of which are contributing factors to conversion to dementia^7,8^. Not surprisingly, hypertension has been identified as one of the 12 modifiable risk factors that account for the approximate 40% of dementia cases that could be delayed or prevented.^1^ Most BP research has examined the detrimental effects of elevated BP; however, older adults can also experience visit-to-visit variability in their BP.

Variability in BP over time has been observed to be associated with increased risk of developing dementia.^9,10^ In cognitively normal adults with a genetic risk (i.e., APOE Ɛ4 positivity) of developing Alzheimer’s disease (AD), BP variability is associated with tau and amyloid alterations^11^ and medial temporal lobe atrophy.^12^ Furthermore, one study following ~13,000 cognitively normal adults for a median of 5 years showed that increased BP variability was associated with increased cognitive decline and vascular pathology (WMHs, atherosclerosis, and infarcts) and AD-related pathology (neurofibrillary tangles).^13^ With respect to AD, BP variability is associated with increased rates of cognitive decline^14^. Despite these findings, limitations still exist in our understanding of how BP influences cognitive change and brain structure.

The current study investigated the relationship between long-term BP status (normal vs. high vs. variable BP), cognition, structural brain changes, and postmortem neuropathology. Our goals were to determine if: 1) variable BP was associated with more cognitive and brain declines than normal and/or high BP, 2) variable BP is associated with increased postmortem neuropathology compared to normal and/or high BP, and 3) findings replicate in a secondary cohort.

## 2. Methods

### 2.1 RADC Research Resource Sharing Hub (RUSH)

Data was used from the RADC Research Resource Sharing Hub (www.radc.rush.edu). Participants provided informed written consent to participate in one of three cohort studies on aging and dementia: 1) Minority Aging Research Study^15^, 2) Rush Alzheimer’s Disease Center African American Clinical Core^16^, or 3) the Rush Memory and Aging Project^17^.

#### 2.1.1 Participants

Participants from RUSH had a baseline age of at least 55 and were either cognitively normal or diagnosed with MCI or dementia. Cognitive status was determined using a three-stage process including computer scoring of cognitive tests, clinical judgment by a neuropsychologist, and diagnostic classification by a clinician based on the National Institute on Aging and the Alzheimer’s Association.^18^ Mean and standard deviation (SD) of systolic BP was computed for each individual, taking into account all their longitudinal timepoints. The SD of the whole sample was then calculated based on individual SDs. These SDs were then used to determine if each participant exhibited normal, high, or variable BP. For normal, their mean BP had to be less than 130 and their SD had to be not more than 1 SD away from the sample SD. For high, their BP must have been greater than or equal to 130 (as per the National Institute of Health and National Institute on Aging guidelines for older adults),^19^ and their SD less than 1 SD away from the sample mean SD. Variable BP were participants whose blood pressure SD was more than 1 SD away from the mean SD of the sample. The sample consisted of a total of 4606 older participants with 32776 time points. There were 1332 older adults with 9145 timepoints who had normal BP, 1377 with 8602 time points who had high BP, and 1897 with 15018 time points with variable BP.

Additional analyses were completed to examine the influence of BP on brain structure with the subset of participants who either had MRI measures from which volumetric measures could be extracted or postmortem neuropathology information. A total of 1846 participants had postmortem neuropathology information (n=486 normal BP, n=473 high BP, and n=886 variable BP). For MRI measures, a total of 820 participants (n=268 normal BP, n=244 high BP, and n=307 variable BP), with 1555 follow-ups were included (n=532 normal BP, n=458 high BP, and n=563 variable BP).

#### 2.1.2 Cognitive Battery

##### Cognitive Assessments

Participants were administered a comprehensive neuropsychological battery comprised of 19 tests assessing episodic memory, semantic memory, working memory, processing speed, and visuospatial ability^20,21^. For detailed information, please see Supplemental material or https://www.radc.rush.edu/.

#### 2.1.3 MRI and post-mortem measures

All MRI and post-mortem measurements were calculated based on standard procedures determined by the RADC Rush researchers and neuropsychologists. T1-weighted (T1w) 3D Magnetization Prepared Rapid Acquisition Gradient Echo (MPRAGE) and T2-weighted 2D Fluid-Attenuated Inversion Recovery (FLAIR) were acquired for structural assessments. T1w images were processed using FreeSurfer. Total GM and WM volumes as well as intracranial volumes (used for normalization) were calculated using the Computational Anatomy Toolbox (CAT)^22^ toolbox from SPM^23^. WMHs were segmented using sysu^24^, a previously validated deep learning based automated WMH segmentation tool.

Methodology used to determine cerebral atherosclerosis, arteriolosclerosis, cerebral amyloid angiopathy severity, and presence of infarcts was determined by the RUSH RADC investigators. Detailed methods are presented in supplementary material.

### 2.2 Alzheimer’s Disease Neuroimaging Initiative (ADNI) Participants

Data used in the preparation of this article were also obtained from the Alzheimer’s Disease Neuroimaging Initiative (ADNI) database (adni.loni.usc.edu). Participants from ADNI had baseline ages between 55 and 90 (see supplemental material or www.adni-info.org for more information).

Participants were included if they had BP measurements from at least two visits and had information for the dependent variables of interest. That is MRIs with ventricle, hippocampal, and entorhinal cortex volume measurments and had at least one of the cognitive tests available, the Alzheimer’s Disease Assessment Scale-13 (ADAS-13) or Functional Activities Questionnaire (FAQ). A total of 2114 participants with 9827 follow-up time points were included. These participants were then divided into one of three groups, normal BP, high BP, or variable BP. Similar to RUSH, the sample SD was then calculated and used to divide participants into the three groups: 1) normal BP, 2) high BP, and 3) variable BP. The sample consisted of 568 older adults with 2348 timepoints who had normal BP, 771 with 2843 time points who had high BP, and 775 with 4636 time points with variable BP.

#### 2.2.1 Structural MRI acquisition and processing

All longitudinal scans were downloaded from the ADNI website (see http://adni.loni.usc.edu/methods/mri-tool/mri-analysis/ for detailed MRI acquisition protocol). T1w scans for each participant were pre-processed through our standard pipeline including noise reduction^25^, intensity inhomogeneity correction^26^, and intensity normalization into range [0-100]. The pre-processed images were then linearly (9 parameters: 3 translation, 3 rotation, and 3 scaling)^27^ registered to the MNI-ICBM152-2009c average ^28^.

#### 2.2.2 WMH measurements

A previously validated WMH segmentation technique was employed to generate participant WMH measurements^29^ (detailed in supplementary material).

#### 2.2.3 Freesurfer Measurements

T1w images were processed using FreeSurfer and quality controlled by the UCSF group, and regional GM and WM volumes were extracted. 1.5T and 3T data were processed with FreeSurfer versions 4.3 and 5.1, respectively, as appropriate.

## 3. Statistical Analysis

Analyses were performed using ‘R’ software version 4.0.5. Linear mixed effects models investigated rates of change differences in the dependent variables across groups (normal, high, variable BP). The dependent variables included rates of change for global cognition, episodic memory, semantic memory, perceptual speed, perceptual orientation, working memory and structural brain changes that were observed over time (WMHs, GM, WM). Baseline age, sex, and baseline diagnosis were included as covariates. The interaction of interest, TimeFromBaseline:Group, examined if change over time differed between Groups (normal, high, and variable BP). Normal BP was used as the reference group, but the models were repeated a second time using variable BP as the reference to observe differences between high vs variable BP. Participant ID was included as a categorical random effect to account for repeated measures of the same participant.

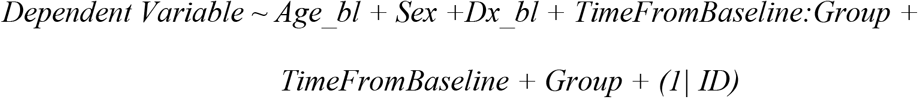

Cerebral atherosclerosis, cerebral amyloid angiopathy, arteriosclerosis, gross chronic cerebral infarcts, chronic microinfarcts assessments were completed post-mortem and were thus analyzed using linear regressions. Age at death, sex, and baseline diagnosis were included as covariates. The effect of interest was group (normal, high, and variable BP), to examine if the dependent variables differed by group.

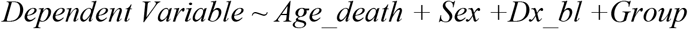

MRI data were not collected at consistent intervals for the RUSH dataset. For example, some participants had MRIs at their baseline visit and then at years 2 and 4, whereas others had MRI information available at years 19 and 21. Therefore, we discarded the information prior to the MRI visits, and considered the first MRI time point as baseline for the MRI analyses and adjusted the *TimeFromBaseline* accordingly. For example, if someone had MRI visits at years 19 and 21, in our MRI analyses, those visits were considered as 0 (baseline) and year 3. All continuous values (except follow-up year) were z-scored within the population prior to analyses.

## 4. Results

Demographic information for both cohorts is presented in table 1. A summary of all results is presented in supplemental materials.

**Table 1:** Demographic Information for both cohorts.

| **Full Sample** | **RUSH** |  | **ADNI** |  |  |  |
| --- | --- | --- | --- | --- | --- | --- |
| Sample Size | 4606 |  | 2114 |  |  |  |
| Age | 76.8 ± 7.7 | | 73.3 ± 7.2 | | | |
| Sex (Female) | 3429 (74%) |  | 1132 (53%) |  |  |  |
| Education | 15.8 ± 4.0 | | 16.0 ± 2.8 | | | |
| BP Group |  |  |  |  |  |  |
| Normal | 1332 (29%) |  | 568 (27%) |  |  |  |
| Variable | 1897 (41%) |  | 775 (37%) |  |  |  |
| High | 1377 (30%) |  | 771 (36%) |  |  |  |
| Diagnosis |  |  |  |  |  |  |
| NC | 3347 (73%) |  | 770 (36%) |  |  |  |
| MCI | 1063 (23%) |  | 977 (46%) |  |  |  |
| Dementia | 200 (4%) |  | 372 (18%) |  |  |  |
| Race |  |  |  |  |  |  |
| Black | 1189 (26%) |  | 95 (5%) |  |  |  |
| White | 3230 (70%) |  | 1948 (92%) |  |  |  |
| Other | 191 (4%) |  | 73 (3%) |  |  |  |
| **Sample by Group** | Rush Normal BP (n=1332) | Rush Variable BP (n=1897) | Rush High BP (n=1377) | ADNI Normal BP (n=568) | ADNI Variable BP (n=775) | ADNI High BP (n=771) |
| Age | 75.7 ± 8.0 | 77.7 ± 7.4 | 76.5 ± 7.7 | 71.6 ± 7.2 | 74.8 ± 7.2 | 73.2 ± 7.0 |
| Sex (female) | 958 (72%) | 1463 (77%) | 1004 (73%) | 264 (46%) | 354 (46%) | 367 (48%) |
| Education | 6.0 ± 4.0 | 15.5 ± 3.8 | 15.9 ± 4.1 | 16.1 ± 2.7 | 16.0 ± 2.8 | 16.0 ± 2.8 |
| Mean BP | 119 ± 7.4 | 137 ± 13.7 | 141 ± 10.2 | 120 ± 9.1 | 139 ± 19.5 | 141 ± 10.9 |
| Diagnosis |  |  |  |  |  |  |
| NC | 983 (74%) | 1348 (71%) | 1013 (73%) | 209 (37%) | 245 (32%) | 314 (41%) |
| MCI | 290 (22%) | 460 (24%) | 313 (23%) | 257 (45%) | 396 (51%) | 321 (42%) |
| Dementia | 59 (4%) | 89 (5%) | 51 (4%) | 102 (18%) | 134 (17%) | 136 (17%) |
| Race |  |  |  |  |  |  |
| Black | 278 (21%) | 529 (28%) | 927 (67%) | 20 (4%) | 29 (4%) | 45 (6%) |
| White | 988 (74%) | 1313 (69%) | 382 (3%) | 529 (93%) | 721 (93%) | 694 (90%) |
| Other | 66 (5%) | 55 (3%) | 68 (5%) | 19 (3%) | 25 (3%) | 32 (4%) |

### 4.1 RUSH

Figure 1 shows the trajectories of cognitive change by BP group over time. Figure 2 shows overall GM, WM, and WMH volume by group.

**Figure 1:**
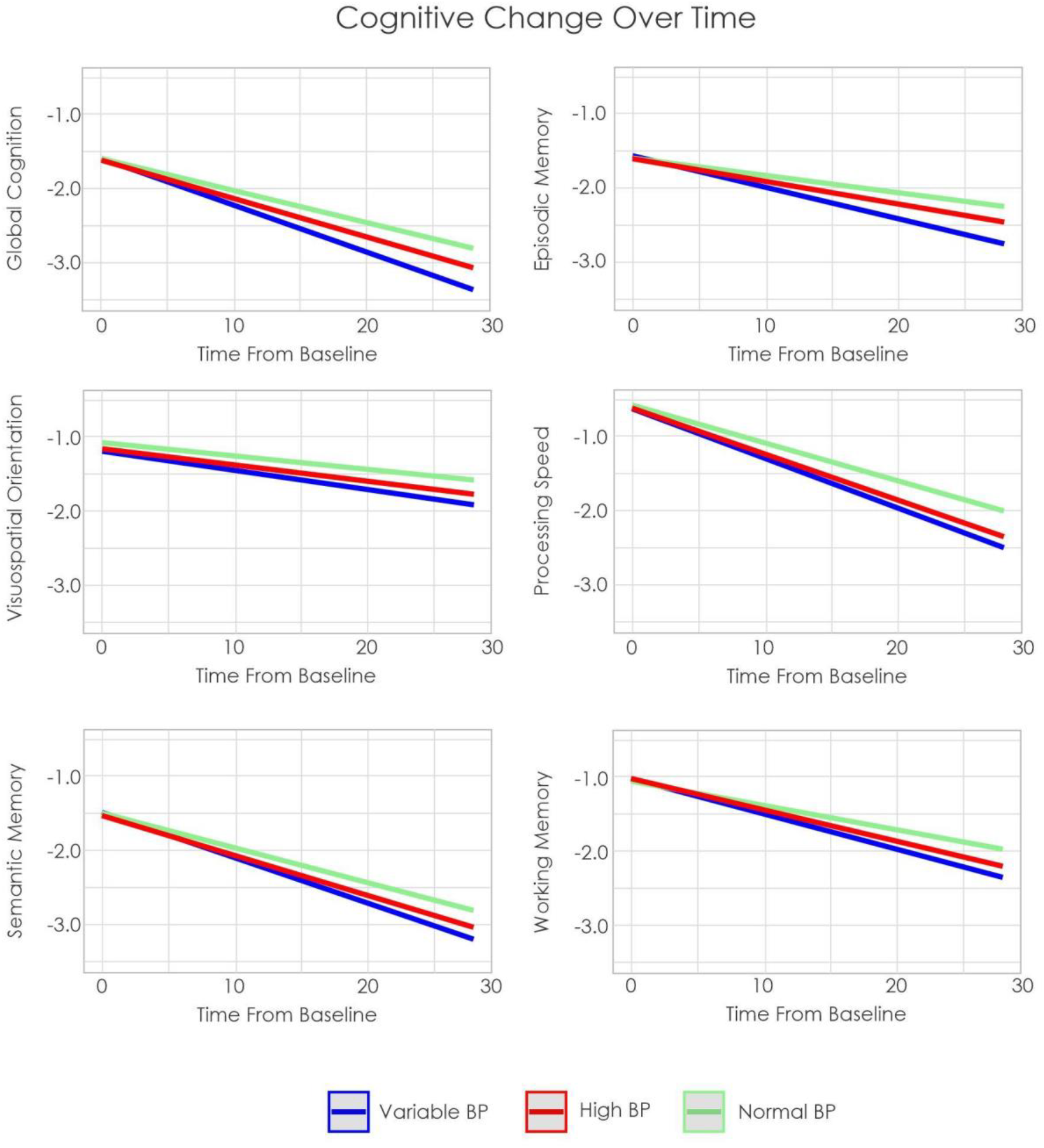
Estimated cognitive change over time by group in RUSH Notes: BP = Blood pressure

**Figure 2:**
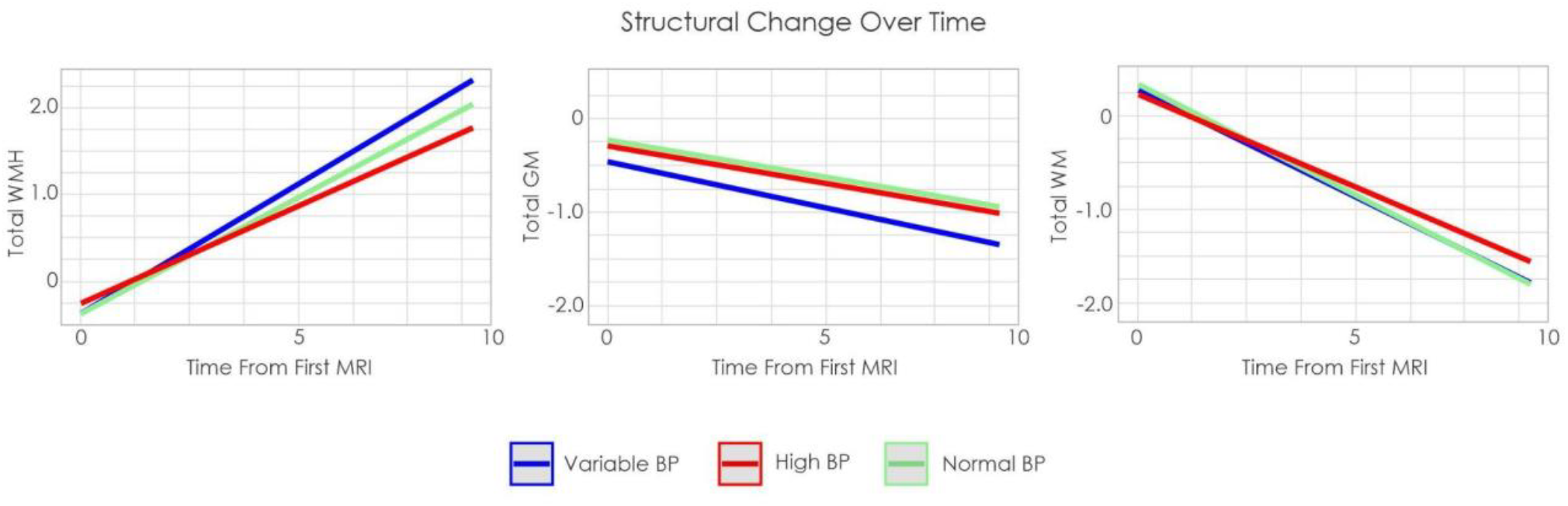
Estimated volume change over time by group in RUSH Notes: WMH = White matter hyperintensity. GM = Gray matter. WM = White matter. BP = Blood pressure

#### 4.1.1 Cognitive Outcomes

Older adults with variable BP had increased rates of decline compared to those with normal and high BP in global cognition, episodic memory, semantic memory, processing speed, and working memory (*t* belongs to [11.57 – 2.69], *p*<.007). For visuospatial orientation, those with variable BP had increased rates of decline compared to those with normal BP (*t*=3.71, *p=*.002), but not high BP (*t*=1.78, *p*=.075). Those with high BP also had increased rates of cognitive decline compared to those with normal BP in global cognition, episodic memory, semantic memory, processing speed, and working memory (*t* belongs to [5.44 – 3.15], *p*<.002). For visuospatial orientation, those with high BP did not differ from normal BP (*t*=1.59, *p*=.11).

#### 4.1.2 MRI Outcomes

Those with normal BP exhibited lower overall WMH burden (*t*= -3.71, *p<*.001) and higher GM volumes (*t*= 2.87, *p=*.003) compared to only those with variable BP. Those with high BP also exhibited lower overall WMH burden (*t*= -3.60, *p<*.001) and higher GM volume than those with variable BP (*t*= 2.59, *p=*.009). Total WM volume slopes did not differ between the three groups.

#### 4.1.3 Post-mortem Outcomes

When examining cerebral atherosclerosis, those with normal BP exhibited less severe ratings than both variable (*t*=-6.37, *p*<.001) and high BP (*t*=-8.16, *p*<.001), and high BP was more severe than variable BP (*t*=2.88, *p*=.004). For arteriosclerosis, those with normal BP had less severe ratings than both variable (*t*=-3.40, *p*<.001) and high BP (*t*=-4.54, *p*<.001), who did not differ. Similarly, for infarctions, normal BP had less severe ratings than both variable (*t*=-4.31, *p*<.001) and high BP (*t*=-3.09, *p*=.002), who did not differ. For microinfarctions, those with normal BP had less severe ratings than only variable BP (*t*=-2.28 *p*=.02). No differences between high BP and either group were observed. There were no significant group differences in cerebral amyloid angiopathy severity.

### 4.2 ADNI

Figure 3a) shows trajectory of cognitive scores by group, figure 3b) shows trajectory of total, cortical, and subcortical GM, and WM volume change over time by group. Figure 4 shows WMH trajectory by group.

**Figure 3:**
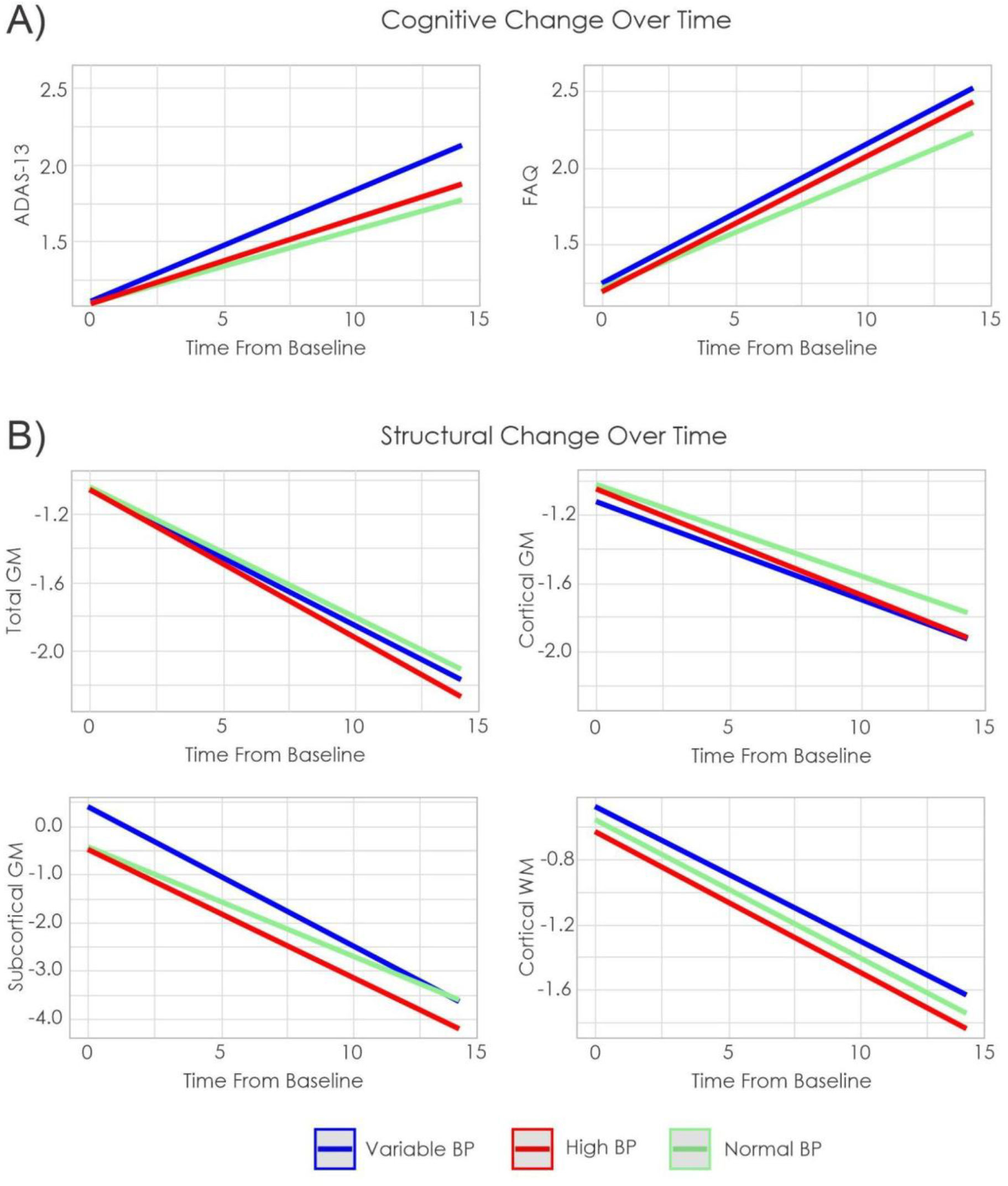
Estimated cognitive and structural brain change over time by group in ADNI Notes: ADAS-13 = Alzheimer’s Disease Assessment Scale-13. FAQ = Functional Activities Questionnaire (FAQ). GM = Gray matter. WM = White matter.

**Figure 4:**
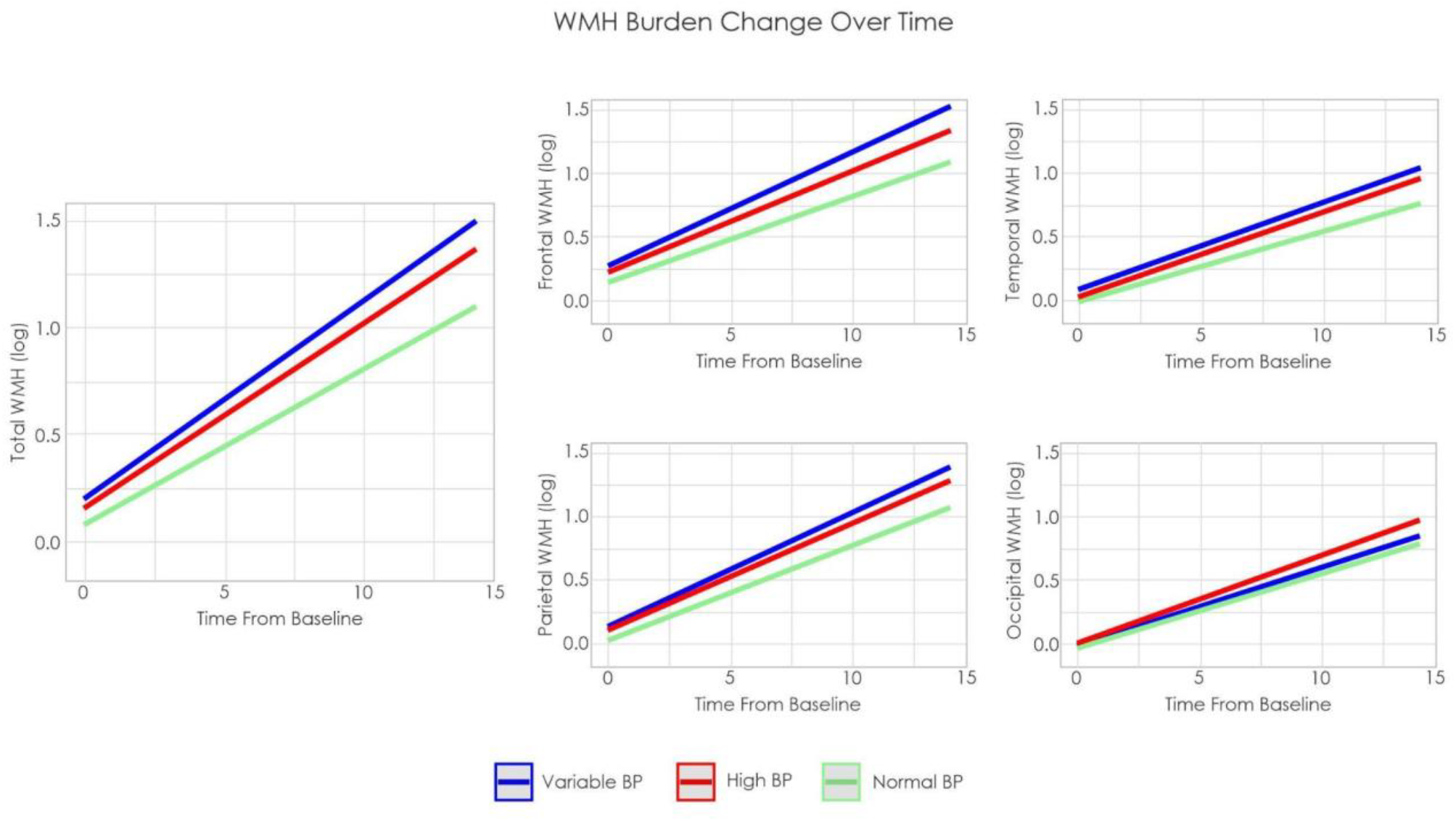
Estimated WMH progression over time by group in ADNI Notes: WMH = White matter hyperintensity. BP = Blood pressure.

#### 4.2.1 Cognitive Outcomes

When looking at global cognition, as measured by the ADAS-13, normal BP had less change than variable BP (*t*= -3.45, *p*<.001), but did not differ from high BP (*t*=0.89, *p*=.37). Furthermore, variable BP had increased cognitive decline compared to high BP (*t*=2.46, *p=*.014). With respect to functional status, as measured by the FAQ, normal BP exhibited greater functional ability compared to variable BP (*t*= -2.56, *p*<.001), but did not differ from high BP (*t*=-1.84, *p*=.03), and high BP did not differ from variable BP (*t*=0.36, *p*=.72).

#### 4.2.2 GM and WM Outcomes

The only difference observed in GM and WM was in subcortical GM. Normal BP exhibited less decline in subcortical GM volume compared to both variable (*t*= -5.19, *p* <.001) and high BP (*t*= -2.85, *p =*.004). Variable BP exhibited increased rate of subcortical volume loss compared to high BP (*t*= 2.47, *p =*.013). No group differences were observed in cortical GM or cortical WM.

#### 4.2.3 WMH Outcomes

When looking at WMH burden change over time, we observed many differences between groups. Normal BP exhibited lower WMH burden increase over time at all regions except occipital compared to both variable (*t* belongs to [-7.50 – -3.80], *p*<.001) and high BP (*t* belongs to [-3.85 – -2.62], *p*<.006). Variable BP exhibited increased total (*t*= 2.18, *p=*.029) and frontal WMH burden compared to high BP (*t*= 3.49, *p*<.004), these groups did not differ at temporal, parietal, or occipital regions. Finally, there were no group differences in the occipital region.

## Discussion

This study examined the relationship between BP and cognition, brain structure, and postmortem neuropathology. The findings show that people with variable and high BP exhibit increased rates of cognitive decline, WMH burden, and vascular pathologies at death. Importantly, those with variable BP exhibited heightened rates of cognitive decline, GM volume loss, and increased WMH burden compared to those with normal and high BP. These findings suggest that while both high and variable BP are detrimental to cognitive decline and structural brain changes, variable BP may experience more negative complications due to their BP fluctuations. Our findings support the established theory that high BP is a modifiable risk factor that contributes to cognitive decline and dementia.^1^ Expanding beyond that understanding, is that variable BP may have more severe implications for cognition and structural brain changes.

Increased cognitive decline was observed in those with high and variable BP compared to older adults with normal BP. Those with variable BP also exhibited increased decline compared to older adults’ high BP. This finding was observed in global cognition, episodic memory, semantic memory, and working memory. When examining visuospatial orientation and functional status, only those with variable BP exhibited increased rates of decline compared to those with normal BP. Although previous studies have reported that BP increases risk for dementia,^9,10^ a detailed examination into the cognitive domains influenced by BP remained unexplored. These findings suggest that variable BP impairs cognitive and functional status above and beyond what occurs due to high BP. These cognitive deficits occur alongside changes in brain structure.

Consistent with previous findings in healthy older adults over a 5-year period,^13^ we observed that BP variability was associated with increased vascular pathology and WMHs. We also observed that high and variable BP was associated with lower GM volume than normal BP. This finding is also consistent with previous research that observed that high BP leads to reductions in brain volume and may be an important factor for neurodegeneration.^30^ Again, the increase rate of change in variable BP observed in both datasets suggests that variable BP may have more detrimental effects on GM volume and overall neurodegeneration than high BP. Increased rates of WMH burden were also observed in high and variable BP groups compared to normal BP except in the occipital region. In the RUSH dataset, we observed that the variable BP group had increased WMHs compared to high and normal BP who did not differ. However, we observed differences between high and normal BP in the ADNI dataset. These differences may be associated with the regional method employed to analyze the ADNI data. Regionally, high and variable BP did not differ from each other in the temporal or parietal regions, but both had more WMH burden than normal BP. However, total and frontal WMH burden was progressively worse from normal to high to variable BP. Previous work has observed that different patterns of WMH accumulation indicates different etiologies.^31–34^ For example, a more widespread distribution of WMHs is associated with non-amnestic MCI^33^ [which leads to dementia other AD] whereas posterior WMHs are strongly associated with conversion to AD.^31^ Frontal WMHs are more strongly associated with vascular risk factors (such as hypertension), indicating that variable BP has more negative effects on brain structure resulting in increased WMH in frontal regions compared to high BP.

With respect to post-mortem neuropathology, we observed that variable and high BP were associated with increased changes compared to normal BP. Both high and variable BP groups had increased amounts of arteriosclerosis and infarctions compared to normal BP but did not differ from each other. Further, variable BP displayed increased microinfarctions compared to normal BP, whereas high BP did not differ from either normal or variable. This finding suggests that variable BP may be more strongly associated with microinfarctions than high BP. These differences in BP variability are also consistent with a previous study that showed increased BP variability was associated with increased arteriosclerosis, infarctions, and microinfarctions^13^. However, they also observed that high variability was associated with increased cerebral amyloid angiopathy which we did not observe. This difference may be associated with study design, as we separated our participants into three groups (normal, high, variable BP) whereas they looked at variability between visits as a continuous measure.

The underlying biological mechanism linking BP variability to atrophy and cognitive decline is largely unknown. Previous work has suggested a role of subclinical cerebrovascular injury as a major pathway linking BP variability and dementia^9^. That is, high levels of BP variability lead to increased cerebral blood flow variability, and consequently to cerebrovascular injury, neurodegeneration, and cognitive decline. Our findings regarding increased rates of WMH accumulation, GM atrophy, and post-mortem vascular pathology in individuals with variable BP provide support for this hypothesis.

This study observed that high and variable BP are associated with increased rates of cognitive decline, neurodegeneration, WMH burden, and post-mortem neuropathology. Variable BP was more strongly associated with increased rate of change than high BP. These declines due to BP may reduce resilience to future pathology and cognitive decline due to dementia. Given that BP can be managed with lifestyle changes and medications, and that no randomized clinical trials have previously considered BP variability as a treatment target, more investigations into management of BP variability as a treatment target for reducing subsequent cognitive decline and dementia is warranted.

## Supporting information

Supplemental Methods and Results

## Data Availability

All data produced are available online at adni.loni.usc.edu and www.radc.rush.edu

## Acknowledgments

Data collection and sharing for this project was funded by the Alzheimer’s Disease Neuroimaging Initiative (ADNI) (National Institutes of Health Grant U01 AG024904) and DOD ADNI (Department of Defense award number W81XWH-12-2-0012). ADNI is funded by the National Institute on Aging, the National Institute of Biomedical Imaging and Bioengineering, and through generous contributions from the following: AbbVie, Alzheimer’s Association; Alzheimer’s Drug Discovery Foundation; Araclon Biotech; BioClinica, Inc.; Biogen; Bristol-Myers Squibb Company; CereSpir, Inc.; Cogstate; Eisai Inc.; Elan Pharmaceuticals, Inc.; Eli Lilly and Company; EuroImmun; F. Hoffmann-La Roche Ltd and its affiliated company Genentech, Inc.; Fujirebio; GE Healthcare; IXICO Ltd.; Janssen Alzheimer Immunotherapy Research & Development, LLC.; Johnson & Johnson Pharmaceutical Research & Development LLC.; Lumosity; Lundbeck; Merck & Co., Inc.; Meso Scale Diagnostics, LLC.; NeuroRx Research; Neurotrack Technologies; Novartis Pharmaceuticals Corporation; Pfizer Inc.; Piramal Imaging; Servier; Takeda Pharmaceutical Company; and Transition Therapeutics. The Canadian Institutes of Health Research is providing funds to support ADNI clinical sites in Canada. Private sector contributions are facilitated by the Foundation for the National Institutes of Health (www.fnih.org). The grantee organization is the Northern California Institute for Research and Education, and the study is coordinated by the Alzheimer’s Therapeutic Research Institute at the University of Southern California. ADNI data are disseminated by the Laboratory for Neuro Imaging at the University of Southern California.

We also want to acknowledge all the MARS, AA Core, and MAP participants. We are also grateful for the hard work from the staff and investigators at the Rush Alzheimer’s Disease Center. To obtain data from MARS, AA Core, and MAP for research use, please visit the RADC Research Resource Sharing Hub (www.radc.rush.edu).

## Consent Statement

Written informed consent was obtained from participants or their study partner.

