## Supplemental Methods and Results for "Beyond Hypertension: Examining Variable Blood Pressure’s Role in Cognition and Brain Structure"

Supplemental Material

**Methods**

*RUSH Cognitive Assessment:*

Episodic memory was assessed using seven tests (immediate and delayed recall of Story A of the Wechsler Memory Scale-Revised; immediate and delayed recall of the East Boston Story; Word List Memory, Recall and Recognition). Semantic memory was assessed using three tests (Verbal Fluency; Boston Naming; Reading Test). Working memory was assessed using three tests (Digit Span forward and backward; Digit Ordering). Processing speed was assessed using four tests (Symbol Digit Modalities Test; Number Comparison; two indices from a modified version of the Stroop Test). Visuospatial ability was assessed using two tests (Line Orientation; Progressive Matrices). Composite scores were created for each domain by converting all individual test scores TO z-scores, using the mean and standard deviation from the combined cohort at baseline. Then, all z-scores were averaged for each respective domain. Additionally, a measure of global cognitive function was computed for each participant by averaging individual performance across all 19 tests.

*RUSH Neuropathology*

Cerebral atherosclerosis rating was completed after postmortem examination of the extent of involvement of each artery and number of arteries involved. Ratings included 0 = no significant atherosclerosis observed, 1 = Small amounts in up to several arteries (typically less than 25% vessel involvement) without significant occlusion, 2 = In up to half of all visualized major arteries, with less than 50% occlusion of any single vessel, and 3 = In more than half of all visualized arteries, and/or more than 75% occlusion of one or more vessels.

Arteriolosclerosis was used to describe histological changes (e.g., include intimal deterioration, smooth muscle degeneration, and fibrohyalinotic thickening of arterioles with consequent narrowing of the vascular lumen) observed in the small vessels. The vessels of the anterior basal ganglia were assessed with a semiquantitative grading system: 0 = none, 1= mild, 2 = moderate, 3 = severe.

Cerebral amyloid angiopathy pathology was determined using a semiquantitative rating in 4 neocortical regions: midfrontal, midtemporal, parietal, and calcarine cortices. Scores were classified into a 4-level severity with ratings determined by neuropathologist: 0 = none, 1= mild, 2 = moderate, 3 = severe.

Presence of one or more gross chronic cerebral infarctions as well as chronic microinfarcts were determined by neuropathologic evaluations performed at Rush, blinded to clinical data, and reviewed by a board-certified neuropathologist. Participants outcomes were reported as, 0 = no gross chronic infarction or 1 = one or more infarctions (regardless of location) and 0 = no chronic infarctions or 1 = one or more chronic microinfarctions (regardless of location).

*ADNI Information*

The ADNI was launched in 2003 as a public-private partnership, led by Principal Investigator Michael W. Weiner, MD. The primary goal of ADNI has been to test whether serial MRI, positron emission tomography (PET), other biological markers, and clinical and neuropsychological assessment can be combined to measure the progression of mild cognitive impairment and early AD. The study received ethical approval from the review boards of all participating institutions. Written informed consent was obtained from participants or their study partner. Participants were selected only from all ADNI Cohorts (ADNI-1, ADNI-GO, ADNI-2 and ADNI-3).

Cognitively healthy older adults exhibited no evidence of memory decline, as measured by the Wechsler Memory Scale and no evidence of impaired global cognition as measured by the Mini Mental Status Examination (MMSE) or Clinical Dementia Rating (CDR). MCI participants scored between 24 and 30 on the MMSE, 0.5 on the CDR, and abnormal scores on the Wechsler Memory Scale. Dementia was defined as participants who had abnormal memory function on the Wechsler Memory Scale, an MMSE score between 20 and 26 and a CDR of 0.5 or 1.0 and a probable AD clinical diagnosis according to the National Institute of Neurological and Communicative Disorders and Stroke and the Alzheimer’s Disease and Related Disorders Association criteria^18^.

*WMH Measurements in ADNI*

This technique has been validated in ADNI in which a library of manual segmentations based on 50 ADNI participants (independent of those studied here) was created. The technique has also been validated in other multi-center studies such as the Parkinson’s Markers Initiative^1^ and the National Alzheimer’s Coordinating Center.^2^ WMHs were automatically segmented using the T1w contrasts, along with a set of location and intensity features obtained from a library of manually segmented scans in combination with a random forest classifier to detect the WMHs in new images.^3,4^ WMH load was defined as the volume of all voxels as WMH in the standard stereotaxic space (in mm^3^) and are thus normalized for head size. The volumes of the WMHs for frontal, parietal, temporal, and occipital lobes as well as the entire brain were calculated based on regional masks from the Hammers atlas.^3,5^ The quality of the registrations and WMH segmentations was visually verified by an experienced rater (author M.D.), blinded to participants diagnostic group.

*Results Summary Table*

Supplemental Table 1: Results Summary Table

|  | **RUSH** | **ADNI** |
| --- | --- | --- |
| **Cognition** | V > H > N in global, episodic memory, semantic memory, processing speed, and working memory decline  V > N in visuospatial orientation decline  H did not differ from N or V | V > H & N in global cognition decline  H did not differ from N  V > N in functional status declines  H did not differ from N or V |
| **GM** | V > H & N in GMV loss  H & N did not differ | V > H > N in subcortical GMV loss  No group differences in cortical GMV |
| **WM** | No group differences in WM | No group differences in WM |
| **WMH** | V > H & N in WMH burden  H & N did not differ | V > H > N in total WMH  V > H > N in frontal WMH  V & H > N in temporal WMH  H & V did not differ  V & H > N in parietal WMH  H & V did not differ  No group differences in occipital WMH |
| **Pathology** | H > V > N in cerebral atherosclerosis  V & H > N in arteriosclerosis  H & V did not differ  V & H > N in infarctions  H & V did not differ  V > N in microinfarctions  H did not differ from N or V  No group differences in cerebral amyloid angiopathy | N/A |

*Notes:* V = variable blood pressure group. H = high blood pressure group. N = normal blood pressure group. GMV = gray matter volume. WM = white matter. WMH = white matter hyperintensities
